# Inflammation PET and plasma neurofilament light predict survival in people with progressive supranuclear palsy

**DOI:** 10.1101/2025.08.27.25334139

**Authors:** Noah L Shapiro, P Simon Jones, Elijah Mak, Kamen A Tsvetanov, Julia Goddard, Davi S Vontobel, Robert Durcan, Leonidas Chouliaras, Tim Fryer, Young T. Hong, Franklin Aigbirhio, Amanda Heslegrave, Nicolai Franzmeier, Matthias Brendel, Henrik Zetterberg, John T O’Brien, James B Rowe, Maura Malpetti

## Abstract

Progressive Supranuclear Palsy (PSP) is a primary tauopathy characterised by atrophy and neuroinflammation of the brainstem, basal ganglia and, to a lesser degree of the cortex. This study investigates the association of regional atrophy (structural MRI), neuroinflammation ([11C]-PK11195 PET), peripheral markers of neurodegeneration (plasma Neurofilament light chain (NfL)), and clinical severity (PSP rating scale - PSPRS) with survival in people with PSP.

Fifty-nine people with PSP underwent longitudinal structural MRI, surviving on average 3.2 years from the first scan (MRI cohort). Sixteen participants (PET cohort) within this cohort underwent cross-sectional [11C]-PK11195 PET and blood sampling for plasma NfL. We applied modality-specific Principal Component Analyses on imaging data and ran partial correlations, multivariate regressions and Bayesian models to evaluate the association between survival and imaging patterns, clinical severity, and plasma NfL.

In the PET-cohort, higher levels of localised inflammation in subcortical regions (rho=-0.49, p=0.02, BF=8.07) and plasma NfL (rho=-0.57, p=0.01, BF=4.63) were associated with shorter survival, while PSPRS scores were not significant predictors of survival. Subcortical atrophy was associated with shorter survival in the larger cohort (r=-0.38, p=0.001; β=-0.66, p=0.001). Spearman’s correlations, multivariate regressions and Bayesian models converged to the same results.

Regional subcortical atrophy is a robust biomarker associated with survival in people with PSP that can be utilised in large-scale clinical trials. TSPO PET and plasma NfL offer promising complementary markers for smaller-scale trials, where they may prove more sensitive than clinical scores or structural MRI alone. By linking neuroinflammation to survival, our results also highlight immunotherapy as a promising avenue for disease-modifying treatment in PSP.

## Introduction

Progressive Supranuclear Palsy (PSP) is a primary tauopathy, presenting with postural instability, supranuclear palsy, akinesia and cognitive change ^1^. The early diagnosis of PSP can be challenging, although clinicopathological correlations are high^1^. Biomarkers of PSP have been sought to improve diagnosis, prognostication, and pathophysiology characterisation of PSP. For example, progressive atrophy in brainstem, basal ganglia and later in the cortex, is identifiable by Magnetic Resonance Imaging (MRI) ^2,3^. Four-repeat tau pathology and synaptic loss associated with PSP are identifiable by positron emission tomography (PET) with second-generation tau ligands and SV2 ligands. Neurofilament light chain (NfL) has emerged as a marker of ‘non-specific’ axonal injury and neurodegeneration ^4^, reflecting its predominant expression in axons ^5^. In people with PSP, NfL levels in plasma and cerebrospinal fluid (CSF) are substantially increased ^6,7^. Longitudinal assessment of plasma NfL has been implemented in clinical trials in people with PSP ^8^ showing associations with clinical progression ^6^.

Beyond tau pathology and neurodegeneration, neuroinflammation with microglial activation and astrogliosis is a key contributor to PSP pathogenesis. Human postmortem studies showed that activated microglia colocalise with neuronal fibrillary inclusions in subcortical and cortical regions ^9,10^. PET has enabled the localisation and quantification of neuroinflammation in PSP. The most commonly used PET ligands aiming to characterise neuroinflammation target the 18 kDa translocator protein (TSPO), which is overexpressed in the outer mitochondrial membrane of microglia ^11^. Several TSPO radioligands have been implemented in neurodegenerative diseases ^12^, including the first-generation [^11^C]-PK11195 ^13^. In PSP, [^11^C]-PK11195 PET shows increased signal in subcortical regions ^14,15^, colocalising with tau pathology ^16^, and predicts clinical decline^17^.

Most studies utilise the Progressive Supranuclear Rating Scale (PSPRS) to track disease progression in people with PSP. The PSPRS is a clinically valid and widely used test to characterise disease severity in people with PSP ^18^. However, some items on the PSPRS are not highly sensitive to change, are affected by ceiling effects ^19^, and need to be complemented by other functional measures of activities of daily living, dysphagia, and cognition to enable a complete assessment of clinical decline. While PSPRS provides a more immediate and quantifiable measure of clinical deterioration, in retrospective studies, survival offers a broader and more objective assessment of disease impact over time.

In this study, our primary outcomes are the association between localised microglial-mediated neuroinflammation (measured by [^11^C]-PK11195) and plasma NfL levels with survival in a small sample size of well-phenotyped people with PSP-Richardson’s syndrome (PSP-RS). Our secondary outcome compares the associations of [^11^C]-PK11195 PET, plasma NfL, structural MRI, or PSPRS scores with survival. Our tertiary outcomes replicate previous findings to confirm whether atrophy patterns in subcortical regions and PSPRS scores are associated with survival in a larger and clinically heterogeneous cohort of participants with PSP.

## Methods

### Participants

To address the primary and secondary outcomes we selected data from sixteen deceased people with PSP-Richardson’s syndrome (PSP-RS) that underwent [^11^C]-PK11195 PET, and MRI, as part of the Neuroimaging of Inflammation in Memory and Related Other Disorders (NIMROD) study ^20^. In addition, PSPRS scores and plasma samples were collected within 4 months of PET scan for quantification of Aβ40, Aβ42, GFAP, NfL, and p-tau181 levels. We refer to these N=16 participants as the “PET cohort”. To conduct the analysis that investigated the association between plasma NfL and survival, we had to exclude one participant who had a PET scan but no plasma samples available.

To address the tertiary outcomes we selected data from N=59 deceased participants (which includes the N=16 participants from the PET cohort) in long-term observational studies that comprised of diverse PSP clinical syndromes, including PSP-Richardson’s syndrome (78.2%), PSP with predominant frontal presentation (16.4%), PSP with progressive gait freezing (3.6 %), and PSP with predominant parkinsonism (1.7 %). Participants with PSP underwent clinical assessment, including PSPRS, baseline and follow-up MRI scans (See Supplementary Figure 1 for the distribution of MRI visits across years). We refer to these N=59 participants as the “MRI cohort”.

For both cohorts participants all met clinical diagnostic MDS-PSP 2017 criteria ^1^ under the care of NHS clinics affiliated with the Cambridge Centre for Parkinson-plus. Trained neurologists administered the PSPRS, who may have seen patients and related assessment as part of their NHS clinical appointments, but were not aware of research data results at the time of the PSPRS assessment. By the time of this study, N=13 had donated their brain to the Cambridge Brain Bank, with a neuropathological diagnostic confirmation of PSP in all of them.

Participants with mental capacity gave their written informed consent to take part in the study. For those who lacked capacity, their participation followed the personal consultee process in accordance with UK law. The research protocols were approved by the National Research Ethics Service’s East of England Cambridge Central Committee, and the UK Administration of Radioactive Substances Advisory Committee.

### Imaging parameters and processing

Sixteen patients underwent [^11^C]-PK11195 PET, using dynamic imaging for 75 minutes, with GE Advance and GE Discovery 690 PET/CT scanners (GE Healthcare, Waukesha, USA). In this sub-group of patients (PET cohort), the interval between [^11^C]-PK11195 and cross-sectional MRI scans had mean and standard deviation (SD) of 3.8±3.1 months

The MRI imaging protocols have been described previously ^20, 21^. In brief, all 59 participants (MRI cohort) underwent T1-weighted (T1w) scans resampled to 1 mm^3^ voxel sizes on a 3T MRI machine on Prisma Fit (32.2%), TrioTim (44.1%), Skyra Fit (11.9%), Verio (5.1%), and Signa PET MR (6.8%) (Siemens, Erlangen, Germany, GE Medical Systems, Milwaukee, Wisconsin, USA) For further information on the scanning parameters refer to supplementary material Table 11. All participants underwent one MRI scan at baseline, and n=55 had further MRI scans at follow up visits (up to 7 visits; < 6.07 years from baseline); for a detailed overview on the visit distributions, refer to supplementary material Figure 1. For a detailed description of the quality checks and processing of MRI and PET, refer to the supplementary material.

For the MRI cohort, the first MRI obtained is referred to as the MRI at baseline, whereas within the PET cohort, the MRI obtained closest to the PET is referred to as the cross-sectional MRI.

### Blood sample collection and processing

Blood samples were obtained by venepuncture and collected in EDTA tubes ^22^, centrifuged to isolate plasma, then aliquoted and stored at −70°C. Plasma assays were conducted at the UK Dementia Research Institute biomarker laboratory. For a detailed description of blood processing, refer to the supplementary material.

### Statistical Analyses

Statistical analyses were performed in R Studio (version 4.3.2). For time intervals between modalities (MRI, PET, PSPRS, blood draw) we fitted a one-way analysis of variance (ANOVA) to quantify whether the mean difference between modalities was significantly different. For the remaining variables, we calculated the mean and standard deviation across the PSP cohort (Table 1).

**Table 1.** Demographic and clinical details of participants with PSP.

|  | MRI full cohort | PET sub-cohort |
| --- | --- | --- |
| N patients | 59 | 16 |
| Sex (F/M) | 28/31 | 7/9 |
| Age at MRI (mean ± SD) | 70 ± 7 | 69 ± 6 |
| Years of education (mean ± SD) | 13 ± 3 | 12 ± 2 |
| Baseline PSPRS score | 27 ± 10 | 42 ± 14 |
| Time from onset to MRI (mean ± SD) | 3.7 ± 2.2 | 5 ± 1.5 |
| Definite PSP at postmortem | 13/13 | 8/8 |

SD = Standard Deviation

First, a linear mixed effect model was applied to the longitudinal regional brain volumes across N=59 to estimate the annualised rate of brain volume changes (MRI-Slopes), after testing for the model assumptions. We allowed the model to estimate subject-specific intercepts and slopes (annual rate of change) where the regional brain volume was predicted by time (in years). Then, subject-specific slopes were extracted and included in further analyses.

Second, for each modality in the MRI cohort (N=59; MRI at Baseline and MRI-Slopes), and in the PET cohort (N=16; cross-sectional MRI and [^11^C]-PK11195 PET), a Principal Component Analysis (PCA) was performed separately to reduce the dimensionality of the four modality-specific datasets. To enhance the interpretability of the resulting components, we flipped the variable- and participant-specific loadings onto the components to fit the neurobiological interpretations: lower values on MR components reflect volume loss, while higher values on TSPO PET components reflect higher inflammation. A Bartletts Test on the four modality-specific datasets confirmed that the variables could be meaningfully reduced in components. We retain components that had eigenvalues > 1 (Kaiser Criterion), and cumulative explained variance ≥ 80%. For each modality, we applied a varimax rotation on all retained components, and extracted subject scores for 2^nd^ level analyses.

Next, we implemented association analyses. As for a priori hypothesis, we focused on the modality-specific component loaded onto PSP-core regions, including brainstem, cerebellum and basal ganglia, and we investigated one-tailed correlation analyses Specifically, we tested whether proxies of neurodegeneration (lower MRI volumes, higher plasma NfL levels), neuroinflammation (higher PET signal), or clinical severity (higher PSPRS scores) were associated with shorter survival.

To address the primary and secondary outcomes in the PET-cohort, we applied partial Spearman correlations to test the association of survival (time from PET scan to death) with (i) subcortical atrophy (volumes on cross-sectional MRI), (ii) subcortical inflammation, as quantified by [^11^C]-PK11195 PET, (iii) plasma NfL levels and (iv) clinical severity (PSPRS score). Each correlation analysis included disease duration (time from onset to PET) as a covariate. Given the small sample size of the cohort, we applied both Frequentist and Bayesian Spearman’s correlations (https://osf.io/gny35/), allowing more robust statistical interpretation by Bayes Factors (BF). Next, we ran linear models on PSP-core components and plasma NfL levels to predict survival, adjusting for sex, age, years of education, and disease duration. Beyond the hypothesis-driven analyses, we ran explorative correlation analyses with the remaining components and survival, adjusting for disease duration. Finally, a Fisher’s Z transformation analysis ^23^ included modality-specific significant rho correlation coefficients to compare associations between modalities and survival. A p-value > 0.05 suggests neither modality is more strongly associated with survival.

To address the tertiary outcomes we tested the prognostic value of baseline MRI and annual rates of change in regional volumes. Across n=59 (MRI cohort) patients with baseline and longitudinal MRI scans, as assumptions for parametric tests were met, we applied Pearson’s correlations between modality-specific components and survival, correcting for disease duration. We further tested the prognostic value of the PSPRS on survival. Each correlation was complemented by linear models, including survival as the outcome variable, the modality-specific PSP-core component as a main predictor (or the PSPRS), and sex, age, years of education, and disease duration as covariates. Beyond the hypothesis-driven analyses, we ran explorative correlation analyses with the remaining imaging components and survival, adjusting for disease duration.

For all correlation analyses, we set the alpha at 0.05 to reject or not reject the null hypothesis and estimated 95% Confidence Intervals (95%CI) of the correlation coefficient. We set a BF ≥ 3 to quantify evidence supporting the alternative hypothesis under the Lee & Wagenmakers (2014) criteria ^24^. In addition, we also highlight effects with a BF ≥ 2 that support “anecdotal” evidence of the alternative hypothesis, indicating preliminary findings that warrant further investigations.

## Results

Table 1 provides demographic summaries and clinical details of patients in each cohort. For the PET cohort, a one-way ANOVA was applied to the time between PET and each of the other predictors considered. The ANOVA indicated that the time lapse between modalities was not statistically significant F(2,44)=1.239, p=0.3. All variables of interest were obtained within 4 months of PET.

### PET Cohort

#### Subcortical inflammation but not cortical inflammation is associated with survival

The PET-specific PCA identified 4 components from [^11^C]-PK11195 regional values (cumulative explained variance 82%). The second component captured subcortical inflammation and was loaded onto PSP-core subcortical regions (Figure 1D). Notably, the first component captured cortical inflammation. For regional loadings onto the remaining components, see Supplementary Material Table 7. Spearman’s partial correlation between survival and the PET PSP-core component (component 2) identified a significant negative association (rho=-0.49, 95%CI [−0.79,0.007], p=0.02, BF=8.07; Figure 2D). In the linear model the component demonstrated a significant negative association (β=-0.96, SE=0.39, t=-2.45, p=0.03, R²=0.40). See supplementary table 4 for explorative correlations with the other components, which did not identify any significant association with survival.

**Figure 1.**
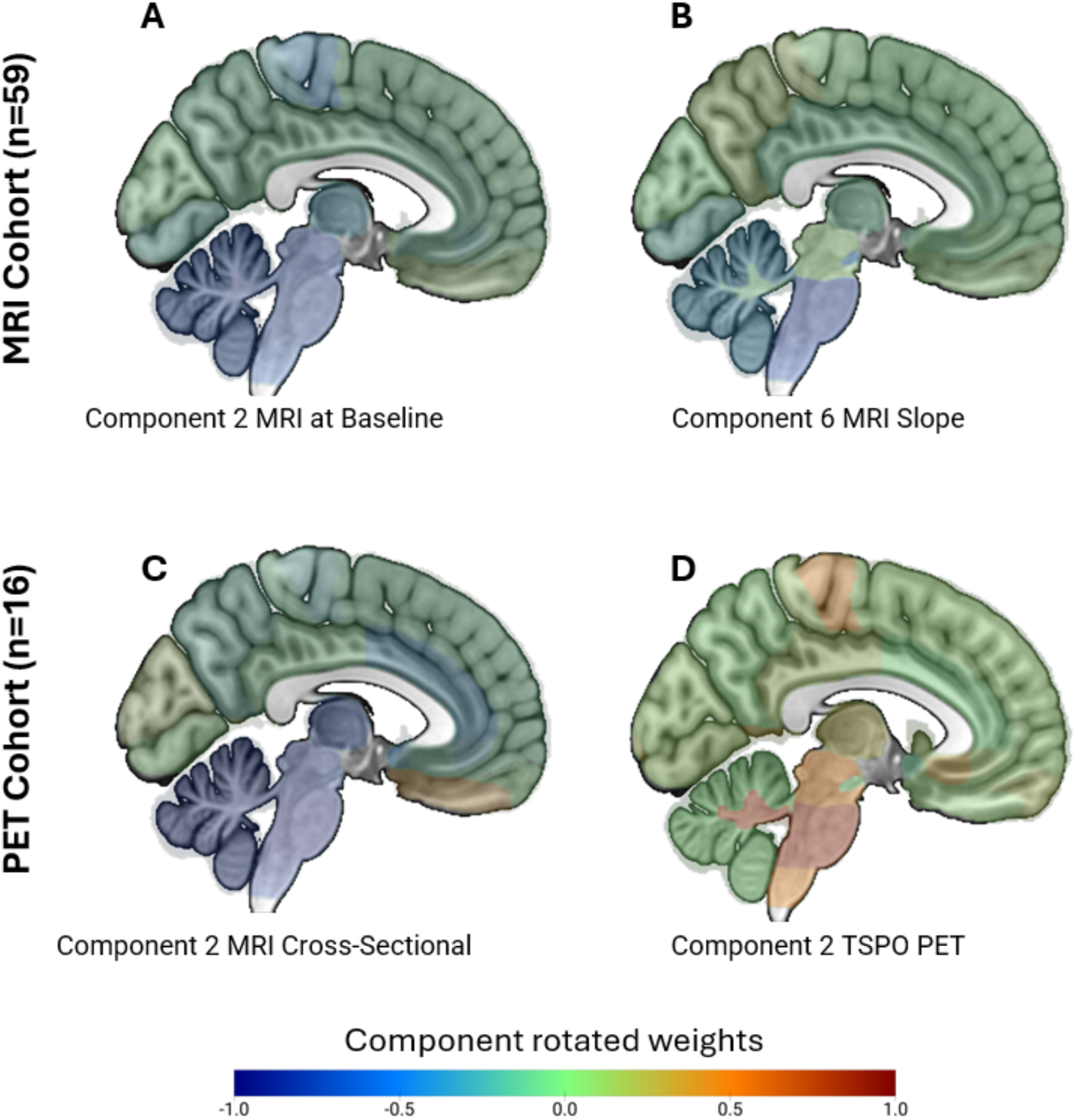
Components reflecting patterns of atrophy and neuroinflammation in PSP core regions. PCA of the MRI cohort identified subcortical components of the baseline MRI volumes (A) and volumetric slopes (annual rate of change) (B). PCA of the PET cohort identified subcortical components in the regional MRI volumes (C) (on the MRI scan closest to PET) and in regional neuroinflammation(D) indexed by [^11^C]-PK1195 (TSPO PET). See Supplementary Tables 7 - 10 for the other significant components. The colour bar indicates regional contributions (correlations) to each component: in MRI components blue regions represent the most atrophic areas, while in the PET component orange/red represent highly inflamed regions.

**Figure 2.**
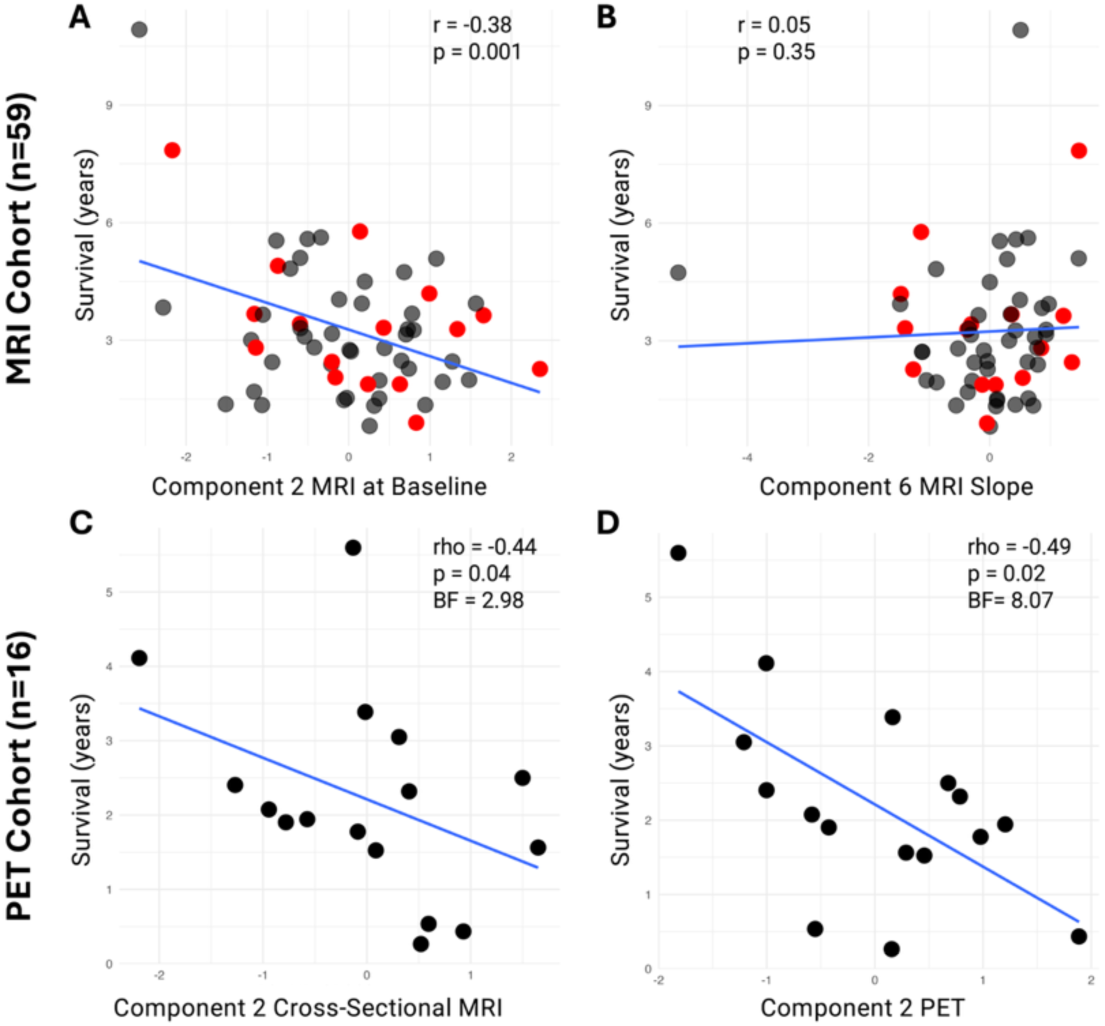
Subcortical atrophy and neuroinflammation predict shorter survival. Scatter plots for the MRI cohort show partial Pearson’s correlations between the hypotheses-driven subcortical and brainstem components (components 2 & 6) of MRI at baseline (A) and for MRI slope (B) with survival, controlled for disease duration. The red dots indicate participants who also are part of the PET cohort. Scatter plots for the PET cohort show partial Spearman’s correlations between the hypotheses-driven subcortical and brainstem components of MRI (C) and of TSPO PET (D) with survival, controlled for disease duration.

#### Plasma NfL is associated with survival

Spearman’s partial correlation between survival and plasma NfL identified a significant negative association (rho=-0.57, 95%CI [−0.83,-0.08], p=0.01, BF=4.63; Figure 3A). In the linear model (R²=0.32, plasma NfL levels were not a statistically significant predictor of survival (β=-4.80, SE=2.43, t=-1.97, p=0.08). See Supplementary Table 6 for exploratory correlations with other plasma biomarkers. Notably, plasma NfL levels significantly correlated with PET PSP-core subcortical component 2 (rho=0.52, 95%CI [0.01,0.81], p=0.02, BF=2.6; Figure 3B) but not with the PET thalamocortical component (component 1; rho=-0.09, 95%CI [−0.57,0.44], p=0.6, BF=0.24).

**Figure 3.**
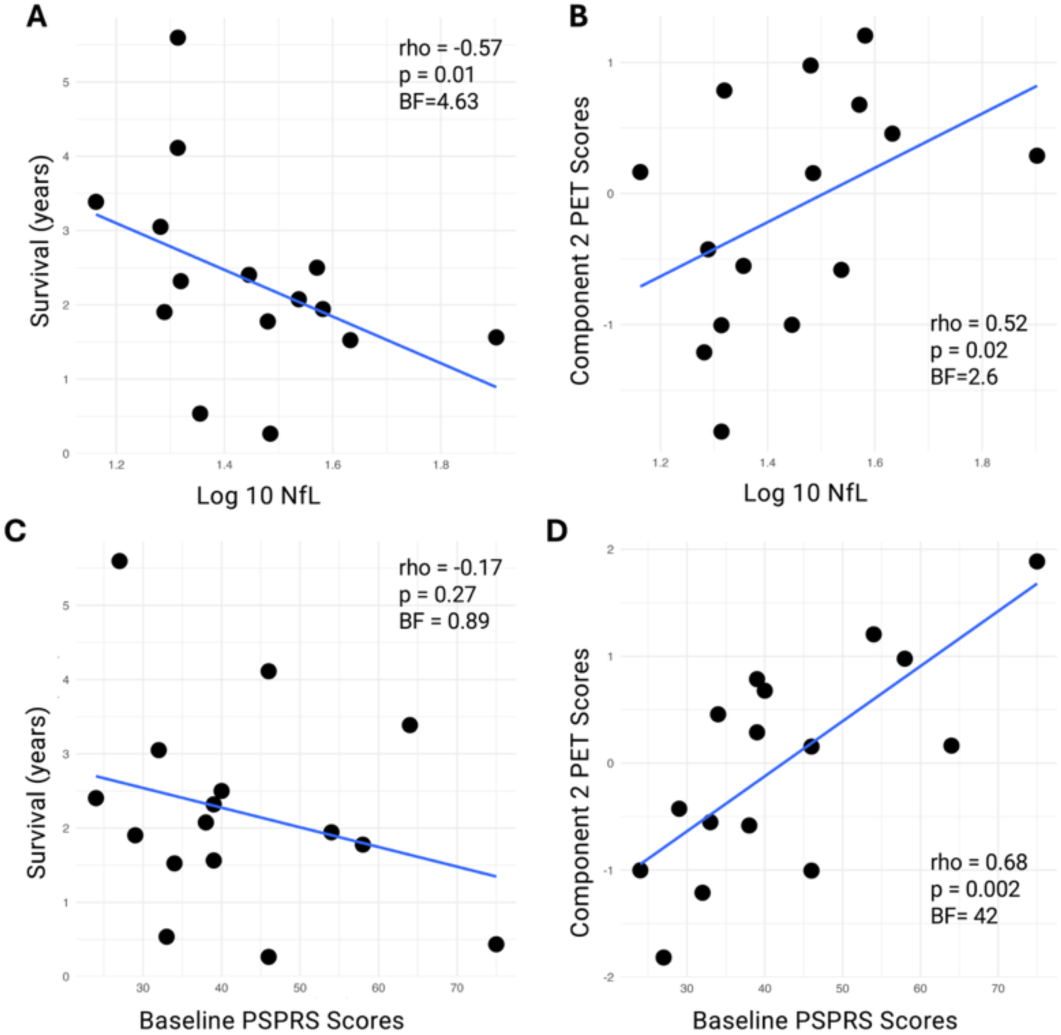
Plasma NfL levels, but not PSPRS scores, correlate with survival in the PET cohort. Scatter plot A shows the partial Spearman’s correlation between the log_1_0-transformed NfL levels and survival, controlled for disease duration. Scatter plot C shows the association between baseline PSPRS scores and survival, controlled for disease duration. Scatter plots B and D represent, respectively, associations of log10-transformed NfL levels and of PSPRS scores with subcortical inflammation.

Including an interaction term between plasma NfL levels and subcortical inflammation did not identify significant interaction effects on survival (NfL*Component2: β=4.97, SE=4.26, t=1.16, p=0.28).

#### The clinical PSP Rating Scale scores is not associated with survival

Spearman’s partial correlation between survival and baseline PSPRS score identified no statistically significant association (rho=-0.17, 95%CI [−0.61,0.33], p=0.27, BF=0.89; Figure 3C). In the linear model, baseline PSPRS scores were not a statistically significant predictor of survival (β=-0.02, SE=0.03, t=-0.625, p=0.5, R^2^=0.09). Notably, the PET PSP-core component (subcortical component 2) significantly correlated with baseline PSPRS (rho=0.68, 95%CI [0.27,0.87], p=0.001, BF=42; Figure 3D). Further baseline PSPRS does not correlate with plasma NfL levels (rho=0.14, 95%CI [−0.40,0.68], p=0.30, BF=0.32).

#### Structural MRI is associated with survival in the PET cohort

In the cohort of patients with PET (n=16), the PCA on cross-sectional MRI identified 6 components (cumulative explained variance 81%). Component 2 captures atrophy in PSP-core regions (Figure 1C). For regional loadings onto the remaining components, see Supplementary Material Table 8. Spearman’s partial correlation between survival rates and the cross-sectional MRI PSP-core component (component 2) identified a significant negative association (rho=-0.44, 95%CI [−0.76,0.07], p=0.04, one-tailed, BF=2.98 (Figure 2C). In the linear model, the component was not a statistically significant predictor of survival (β=-0.61, SE=0.44, t=-1.39, p=0.19, R²=0.20). See Supplementary Table 5 for exploratory correlations with the other components.

#### Comparisons of MRI, PET and plasma NfL to predict survival

Fisher’s Z test compared the rho correlation coefficients of PET-survival (rho_PET_=-0.49, n_PET_=16) and NfL-survival (rho_NfL_=-0.57, n_NfL_=15) associations, suggesting no significant differences (z=0.27, p=0.78). The Fisher’s Z test also compared MRI-survival (rho_MRI_=-0.46, n_MRI_=16) and PET-survival (rho_PET_=-0.49, n_PET_=16) associations, showing no statistically significant differences (z=-0.09, p=0.92). Lastly, the Fisher’s Z test compared the rho correlation coefficients of MRI-survival (rho_MRI_=-0.46, n_MRI_=16) and NfL-survival (rho_NfL_=-0.57, n_NfL_=15) associations, showing no statistically significant differences (z=0.37, p=0.70).

### MRI Cohort

#### Structural MRI is associated with survival in the MRI cohort

Consistent with the established literature, we found that most ROIs showed significant volume loss over time (Supplementary Material Table 1 and Figure 2). The PCA on regional MRI volumes at baseline identified 10 significant components (cumulative explained variance 81%). Component 2 captures atrophy in PSP-core regions (Figure 1A). For regional loadings onto the remaining components, see Supplementary material Table 10. The PCA on regional annual rates of change (slopes) also identified 10 significant components (cumulative explained variance of 80%). Component 6 captures the rate of brain volume loss in PSP-core regions (Figure 1B). For regional loadings onto the remaining components, see Supplementary material Table 9.

The Pearson partial correlation between survival and the baseline-MRI PSP-core component (component 2) showed a significant negative association (r=-0.38, 95%CI [−0.57,-0.13], p=0.001; Figure 2A). In the linear model the component showed a significant negative predictive effect (β=-0.66, SE=0.20, t=-3.282, p=0.001, R^2^=0.32) alongside years of education (β=-0.17, SE=0.06, t=-2.67, p=0.01). See Supplementary Table 2 for explorative correlations with the other components. The Pearson partial correlation between survival and MRI-slope PSP-core component (component 6) showed no significant negative association (r=0.05, 95%CI [−0.20,0.30], p=0.35; Figure 2B). In the linear model the component was not a statistically significant predictor of survival (β=0.15, SE=0.21, t=0.73, p=0.46, R^2^=0.19). See Supplementary Table 3 for exploratory correlations with the other components.

#### The clinical PSP Rating Scale scores is not associated with survival in the MRI cohort

The Pearson partial correlation between survival and the PSPRS showed a significant negative correlation (r=-0.25, 95%CI [−0.47,0.001], p=0.02). In the linear model however the PSPRS showed no significant predictive effect (β=-0.05, SE=0.02, t=-1.978, p=0.05, R^2^=0.16) on survival where years of education was a better predictor of survival (β=-0.17, SE=0.07, t=-2.53, p=0.01).

## Discussion

Our study found that higher subcortical neuroinflammation, quantified by [^11^C]-PK11195, and neurodegeneration, reflected by plasma NfL levels, are both associated with shorter survival in people with PSP. However, significant interaction effects between plasma NfL and subcortical inflammation on survival were not identified, with Frequentist and Bayesian approaches converging to the same conclusion. In the larger cohort, we also replicated previous findings showing that regional atrophy in subcortical regions is associated with survival. Regional subcortical atrophy is a robust prognostic biomarker that can be utilised in large-scale clinical trials, while TSPO PET and plasma NfL may offer complementary and more sensitive information for smaller-scale trials.

Neuroinflammation has been previously described as part of the pathological changes associated with PSP. Specifically, postmortem and TSPO PET studies showed that neuroinflammation co-localises and parallels with the progression of tau pathology from subcortical to cortical regions ^25,16,26^. Inflammation in PSP-core subcortical regions has also been shown to predict faster clinical progression in people with PSP, considering longitudinal PSPRS scores, which captures worsening of symptoms ^17^. Our study expanded on the previous evidence, identifying that subcortical-localised inflammation is also associated with shorter survival in people with PSP. We used survival, instead of longitudinal PSPRS to capture a definitive and objective aspect of prognosis. Considering survival as a clinical outcome, previous studies found peripheral markers of neuroinflammation being clinically relevant and significant prognostic measures in PSP and related conditions. High serum levels of pro-inflammatory cytokines, associated with neuroinflammation, have been linked with shorter survival in patients on the frontotemporal lobar degeneration spectrum ^27^. The association between TSPO PET and inflammatory cytokines allows us to disentangle central inflammatory processes and its role in accelerated disease progression. In PSP, higher numbers of classical monocytes and natural killer cells, associated with lower numbers of TREM2+ cells, were associated with shorter survival ^28^. Furthermore, genetic variations at the Leucine-rich repeat kinase 2 (LRRK2) locus are also associated with survival in PSP ^29^, highlighting the mechanistic contribution of immune pathways to PSP progression. LRRK2 lowering therapies have been proposed in clinical trials for other neurogenerative diseases ^30^, however, the link between LRRK2 levels and regional *in-vivo* brain inflammation in PSP remains to be elucidated. Future research may clarify the relationships between peripheral markers of inflammation, LRRK2 and brain inflammation, and whether patients with PSP might benefit from LRKK2-lowering or cytokine-targeting therapies.

Plasma NfL is a marker of axonal injury and proxy of neurodegeneration, which is commonly used as a biomarker in clinical trials to monitor drug efficacy and disease progression in diverse neurodegenerative disorders ^31,32,33^. Plasma NfL relates to brain hypometabolism, atrophy over time, and clinical progression. In our small cohort, high levels of plasma NfL were associated with shorter survival in people with PSP, aligning with similar findings in larger cohorts ^34,6^. Conversely, GFAP levels were not significantly associated with survival in people with PSP (supplementary material Table 6). Previous research also suggests that GFAP may be a sensitive marker for astrogliosis related to amyloidosis in AD ^35^, but less useful in non-AD pathologies, including PSP ^22^. This, paired with the lack of statistically significant associations between PSPRS scores and survival, suggests that clinical trials using NfL as an outcome of interest may achieve meaningful interpretation of the results with smaller sample sizes, at least at a group level. Interestingly, in our cohort, plasma NfL levels were strongly associated with neuroinflammation in PSP-core subcortical regions, indicating that those patients with higher NfL levels have higher localised brain inflammation. Contrary, plasma NfL levels did not correlate with widespread inflammation in cortical areas. This suggests that NfL levels can inform regional specificity of other PSP-core brain changes and provide an accessible predictor of survival on a group level that may be used in future clinical trials. Moreover, plasma NfL may be a better than other clinically validated markers for patient stratification in relatively small clinical trials. We suggest that NfL levels should be evaluated in addition to clinical and inflammatory markers in clinical evaluation and prediction of clinical deterioration. To date, no other studies including patients with PSP have investigated the association between NfL and TSPO radiotracers. Thus, further studies are needed to expand and validate our results in other cohorts and with different TSPO tracers.

Beyond plasma NfL, a commonly and clinically used measure of neurodegeneration is structural MRI. Large studies have shown that both cross-sectional and longitudinal MRI volumes in subcortical and brainstem regions are associated with survival in people with PSP ^36,37^. In our MRI cohort (n=59), we replicated this result, showing subcortical atrophy at baseline predicts survival in people with PSP. However, in the PET cohort (n=16), we have no statistically significant associations of regional MRI volumetric measures with survival. Similarly, the PSPRS is commonly used in clinics to track clinical severity ^18^ and has been previously shown to be predictive of disease progression and survival. In a cohort of 197 people with PSP, higher scores on the PSPRS predicted a shorter life expectancy ^38^. However, in our PET cohort, baseline PSPRS was not significantly associated with survival, although the directionality of the association was as expected. We suggest that the lack of a statistically significant associations of structural MRI and PSPRS with survival in the PET cohort may be due to lower sensitivity of these measures, in the small sample size, leading to underpowered analyses. On the contrary, our results suggest that TSPO PET signal in PSP-core regions and plasma NfL may be more sensitive and recommendable markers in small-scale clinical trials.

Interestingly, in the MRI cohort (n=59), when baseline MRI was included in the predictive model alongside sex, age, time between onset and MRI scan, and years of education, the latter variable was significantly and negatively correlated with survival. Hence, patients with more years of formal education had a shorter life expectancy. This finding results from initial cognitive and brain reserve as described in other neurodegenerative diseases ^39,40^. For example, in Alzheimer’s disease, higher education as a proxy of cognitive reserve is associated with milder clinical symptomology at a comparable level of brain pathology or worse brain pathology at comparable clinical severity. Thus, patients with higher cognitive reserve are more resilient to brain pathology, showing milder symptomatology, but once pathology passes a critical threshold, individuals with former cognitive reserve experience a more rapid decline. As there is limited data on cognitive reserve in PSP, future research in this field could explore composite scores of reserve, such as occupational attainment, leisure and physical activities, bi or multilingualism, or pre-morbid intelligence quotient ^39^, which might reveal mitigating factors throughout the natural history of the disease.

There are several limitations to our study. First, we acknowledge the small sample size of the PET cohort, which does not permit statistically elaborate models. However, our results found consensus across different analytic methods, including Spearman correlations and linear regression models. The Bayesian tests confirmed that we had sufficient precision (analogous to power in frequentist tests) to support the alternative hypotheses with strong evidence (BF > 3) from the 15 patients’ data. The convergence over these statistical approaches mitigates against inadequate power and sample-dependent biases on the estimation of biomarker–survival associations. Although our cohort is larger than or of comparable size of previous PET studies on rare neurodegenerative diseases like PSP, the replication of these findings with larger and multicenter clinical cohorts will represent an important step to establish the generalizability of our results, and utility for clinical trials. Second, there is a current debate regarding the specificity of [^11^C]-PK11195 PET in quantifying microglia ^41,42^, over and above astrocytes or vascular endothelium ^43^. However, in people with PSP, antemortem regional [^11^C]-PK11195 binding is positively correlated with post-mortem phagocytic microglia and microglial TSPO, providing direct evidence that [^11^C]-PK11195 reflects microglial-mediated brain inflammation ^11^. Aligning evidence were described by Garland et al (2024), who found TSPO being overexpressed and related to phagocytotic microglia in postmortem tissues from patients with Alzheimer’s disease^44^. The [^11^C]PK11195 tracer has limitations, including its relatively low signal-to-noise ratio and low brain penetration which may affect its sensitivity to visualise and quantify microglia. Nevertheless, this would reduce effect sizes and increase type II errors, rather than leading to false positive findings. Second-generation PET radioligands for TSPO are characterised by higher signal-noise ratio than [^11^C]PK11195 but their binding is markedly affected by single nucleotide polymorphisms (rs6971) which cause heterogeneity in PET data and require genetic screening, and lead to the exclusion of participants who are low affinity binders. [^11^C]PK11195 binding is less affected by this polymorphism, has well established methods of kinetic analysis ^45^ and has been validated in PSP with PET-to-postmortem evidence ^11^. Hence, [11C]PK11195 PET was the ligand of choice for this study of PSP. Second, we acknowledge the limited power of the analyses related to the small size of our PET samples. Third, we recruited according to clinical diagnostic criteria, and although clinicopathological correlations of PSP-Richardson’s syndrome are very high, including 8 of 8 cases in our PET-cohort with postmortem pathology, they are not perfect. Finally, our results may not be generalisable to other cohorts and primary tauopathies. For example, in a cohort of people with amyloid-negative corticobasal syndrome (CBS), a recent TSPO PET using the [^18^F]-GE-180 ligand reported that higher brain inflammation was associated with slower clinical progression, as assessed by longitudinal PSPRS scores^46^. The discrepancy with our results may be due to several factors, including (i) regional differences, where PSP is characterised by more localised subcortical inflammation while CBS by widespread cortical inflammation; and (ii) differences in disease severity across cohorts, with previous studies in PSP recruiting on average more advanced participants than in CBS cohorts. Further studies are needed to validate previous evidence across cohorts and diseases.

In conclusion, regional subcortical atrophy is a robust biomarker associated with survival in people with PSP that can be utilised in large-scale clinical trials. TSPO PET and plasma NfL may be more suitable biomarkers for small-scale trials than PSPRS or structural MRI. We further provide evidence that immunotherapeutic strategies are promising avenues to slow disease progression in people with PSP.

## Data Availability

Anonymised data may be shared by request to the senior author from a qualified investigator for non-commercial use (data sharing may be subject to restrictions according to consent and data protection legislation).

## Acknowledgments

We thank our patients with PSP and their families for their participation in this study and the support team who made this research study possible. We thank the National Institute for Health Research (NIHR) Cambridge BioResource centre staff, and the research nurses for their contribution, and the East Anglia Dementias and Neurodegenerative Diseases Research Network (DeNDRoN) for help with subject recruitment.

This study was co-funded by Race Against Dementia Alzheimer’s Research UK (ARUK-RADF2021A-010); the Progressive Supranuclear Palsy Association (G124028); the Dementias Platform UK and Medical Research Council (MC_UU_00030/14; MR/T033371/1); the Wellcome trust (103838; 220258); the Cambridge University Centre for Parkinson-Plus (RG95450); the National Institute for Health Research (NIHR) Cambridge Biomedical Research Centre (BRC-1215-20014; NIHR203312: the views expressed are those of the authors and not necessarily those of the NIHR or the Department of Health and Social Care). This work is also supported by the UK Dementia Research Institute through UK DRI Ltd, principally funded by the Medical Research Council. K.A.T. was funded by a Fellowship award from the Alzheimer’s Society, UK (Grant Number 602). HZ is a Wallenberg Scholar and a Distinguished Professor at the Swedish Research Council supported by grants from the Swedish Research Council (#2023-00356; #2022-01018 and #2019-02397), the European Union’s Horizon Europe research and innovation programme under grant agreement No 101053962, and Swedish State Support for Clinical Research (#ALFGBG-71320). MB was funded by the Deutsche Forschungsgemeinschaft (DFG, German Research Foundation) in a TSPO research unit (ID 403161218) and under Germany’s Excellence Strategy within the framework of the Munich Cluster for Systems Neurology (EXC 2145 SyNergy – ID 390857198). N.L.S was supported by the Cambridge Trust and Sidney Sussex College, University of Cambridge.

## Potential Conflicts of Interest

JTO has received honoraria for work as DSMB chair or member for TauRx, Axon, Eisai and Novo Nordisk, and has acted as a consultant for Biogen and Roche, and has received research support from Alliance Medical and Merck. JBR is a non-remunerated trustee of the Guarantors of Brain, Darwin College and the PSP Association (UK). He provides consultancy unrelated to the current work to Asceneuron, Astronautx, Astex, Curasen, CumulusNeuro, Wave, SVHealth, and has research grants from AZ-Medimmune, Janssen, and Lilly as industry partners in the Dementias Platform UK. M.M. has acted as a consultant for Astex Pharmaceuticals. H.Z. has served at scientific advisory boards and/or as a consultant for Abbvie, Acumen, Alector, Alzinova, ALZPath, Amylyx, Annexon, Apellis, Artery Therapeutics, AZTherapies, Cognito Therapeutics, CogRx, Denali, Eisai, LabCorp, Merry Life, Nervgen, Novo Nordisk, Optoceutics, Passage Bio, Pinteon Therapeutics, Prothena, Red Abbey Labs, reMYND, Roche, Samumed, Siemens Healthineers, Triplet Therapeutics, and Wave, has given lectures in symposia sponsored by Alzecure, Biogen, Cellectricon, Fujirebio, Lilly, Novo Nordisk, and Roche, and is a co-founder of Brain Biomarker Solutions in Gothenburg AB (BBS), which is a part of the GU Ventures Incubator Program (outside submitted work). MB received speaker honoraria from Roche, Iba, GE healthcare and Life Molecular Imaging, is an active advisor of MIAC, and advised GE Healthcare Life Molecular Imaging. NF received speaker honoraria from GE Healthcare, EISAI and Life Molecular Imaging and received research support from Eli Lilly

## Notes

### Author Declarations

Participants with mental capacity gave their written informed consent to take part in the study. For those who lacked capacity, their participation followed the personal consultee process in accordance with UK law. The research protocols were approved by the National Research Ethics Services East of England Cambridge Central Committee, and the UK Administration of Radioactive Substances Advisory Committee.

