## Supplementary material for "Inflammation PET and plasma neurofilament light predict survival in people with progressive supranuclear palsy": Supplementary_Material.pdf

### Blood sample processing

Plasma samples were thawed on wet ice, centrifuged at 500× g for 5 min at 4°C. Calibrators (neat) and samples plasma: 1:4 dilution) were measured in duplicates. The plasma assays used the Quanterix Simoa Human Neurology 4-Plex E assay (measuring Aβ40, Aβ42, GFAP and NfL ) and the Quanterix Simoa p-tau181 measuring p-tau181 of the human tau protein. Assays were performed using the Simoa-HD1 according to the manufacturer's protocol (Quanterix Corp, Billerica, Massachusetts, USA) (Rissin et al). All samples were analysed at the same time using the same batch of reagents. A four-parameter logistic curve data reduction method was used to generate a calibration curve. Two control samples of known concentration of the protein of interest (high- control and low- control) were included as quality controls. All variables were $\log_{10}$-transformed prior to statistical analyses.

### Magnetic Resonance Imaging

We visually inspected all 172 MRI scans to ensure high-quality data; then we resampled all scans to 1 $mm^3$ isotropic resolution using the Advanced Normalization Tools (ANTS) [41] version 2.3.5 function ResampleImage with a Gaussian Interpolation approach. Images were cropped to remove the neck using an in-house script running under MATLAB (MathWorks, inc. Natick, Massachusetts, USA) version 2020b. We segmented the scans into grey matter (GM), white matter (WM), cerebrospinal fluid (CSF), and estimated total intracranial volume (TIV=GM+WM+CSF) using the longitudinal analysis pipeline from the Computational Anatomy Toolbox 12 (CAT12) and statistical parametric mapping software (SPM12). We applied a modified version of the n30r83 Hammersmith atlas (www.brain-development.org), as previously used [16,18,19]. The modified Hammers atlas includes brainstem and cerebellar parcellations, as core regions for PSP, among 83 cortical and subcortical regions of interest (ROI) in every patient. As the scans were obtained with different MRI scanners and protocols, which might introduce noise and bias that could affect subsequent statistical analysis and interpretation, we applied a longitudinal harmonisation algorithm (Long Combat

[42]). All ROIs were then adjusted for TIV, and the left and right corresponding ROIs were summed, resulting in 43 bilateral regional volumes adjusted for TIV.

**Positron Emission Tomography**

For each participant with [$^{11}$C]-PK11195 PET, the aligned dynamic PET image series for each scan was rigidly co-registered to the T1w MRI image. Non-displaceable binding potentials ($BP_{ND}$) were calculated in the same 83 cortical and subcortical ROIs as for T1w MRI. Before kinetic modelling, regional PET data were corrected for partial volume effects from CSF by dividing by the mean regional GM plus WM fraction determined from SPM segmentation. For [$^{11}$C]-PK11195, supervised cluster analysis was used to determine the reference tissue time-activity curve and $BP_{ND}$ values were calculated in each ROI using a simplified reference tissue model with vascular binding correction [43]. The CSF-partial-volume corrected $BP_{ND}$ regional values were averaged across left and right hemisphere, resulting in the same 43 bilateral ROIs as in MRsI processing.

Figure 1

Box plots of MRI visits across years in the MRI cohort. Red lines indicate patients who are also in the PET cohort.

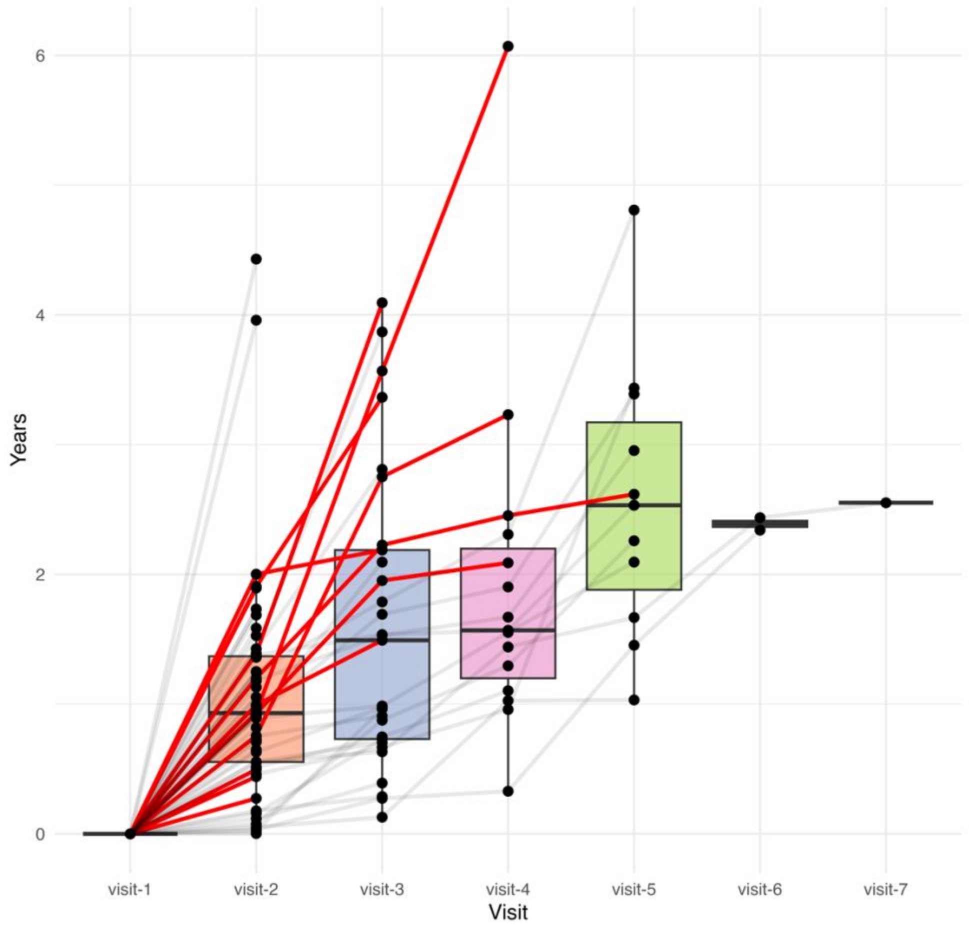

Table 1
An Analysis of Variance (ANOVA) Type 3 Satterthwaite's method was performed on the resulting linear-fixed effect models to determine the effect of time on regional volume for each ROI.

| Regions | Sum Sq | Mean Sq | NumDF | DenDF | F value | Pr(>F) |
|---|---|---|---|---|---|---|
| Brainstem_mid | 0.850 | 0.850 | 1 | 13.233 | 21.359 | 0.000 |
| Brainstem_pon | 0.473 | 0.473 | 1 | 33.095 | 4.179 | 0.049 |
| Brainstem_med | 0.098 | 0.098 | 1 | 39.745 | 15.972 | 0.000 |
| Hippocampus | 0.194 | 0.194 | 1 | 20.683 | 35.725 | 0.000 |
| Amygdala | 0.076 | 0.076 | 1 | 11.100 | 7.256 | 0.021 |
| Anterior_temporal_lobe_medial_part | 0.985 | 0.985 | 1 | 17.820 | 27.031 | 0.000 |
| Anterior_temporal_lobe_lateral_part | 0.243 | 0.243 | 1 | 18.797 | 18.907 | 0.000 |
| Parahippocampal_and_ambient_gyri | 0.085 | 0.085 | 1 | 15.046 | 2.354 | 0.146 |
| Superior_temporal_gyrus_posterior_part | 2.743 | 2.743 | 1 | 20.406 | 39.258 | 0.000 |
| Middle_and_inferior_temporal_gyrus | 6.129 | 6.129 | 1 | 13.251 | 33.681 | 0.000 |
| Fusiform_gyrus | 0.396 | 0.396 | 1 | 19.332 | 18.363 | 0.000 |
| Insula | 3.221 | 3.221 | 1 | 32.762 | 41.471 | 0.000 |
| Lateral_remainder_of_occipital_lobe | 1.316 | 1.316 | 1 | 17.243 | 1.389 | 0.255 |
| Cingulate_gyrus_anterior_part | 1.301 | 1.301 | 1 | 14.012 | 25.276 | 0.000 |
| Gyrus_cinguli_posterior_part | 0.159 | 0.159 | 1 | 15.552 | 2.817 | 0.113 |
| Middle_frontal_gyrus | 32.489 | 32.489 | 1 | 33.929 | 25.846 | 0.000 |

| Regions | Sum Sq | Mean Sq | NumDF | DenDF | F value | Pr(>F) |
|---|---|---|---|---|---|---|
| Posterior_temporal_lobe | 9.798 | 9.798 | 1 | 33.753 | 14.401 | 0.001 |
| Inferiolateral_remainder_of_parietal_lobe | 9.361 | 9.361 | 1 | 40.356 | 21.056 | 0.000 |
| Caudate_nucleus | 0.001 | 0.001 | 1 | 38.455 | 0.026 | 0.873 |
| Nucleus_accumbens | 0.003 | 0.003 | 1 | 24.153 | 8.032 | 0.009 |
| Putamen | 1.488 | 1.488 | 1 | 28.071 | 23.750 | 0.000 |
| Thalamus | 13.459 | 13.459 | 1 | 30.217 | 58.281 | 0.000 |
| Pallidum | 0.009 | 0.009 | 1 | 19.578 | 2.317 | 0.144 |
| Precentral_gyrus | 13.426 | 13.426 | 1 | 24.618 | 27.564 | 0.000 |
| Straight_gyrus | 0.217 | 0.217 | 1 | 18.000 | 10.783 | 0.004 |
| Anterior_orbital_gyrus | 0.854 | 0.854 | 1 | 22.922 | 19.498 | 0.000 |
| Inferior_frontal_gyrus | 4.371 | 4.371 | 1 | 35.543 | 38.282 | 0.000 |
| Superior_frontal_gyrus | 33.154 | 33.154 | 1 | 20.487 | 23.056 | 0.000 |
| Postcentral_gyrus | 6.738 | 6.738 | 1 | 17.149 | 15.143 | 0.001 |
| Superior_parietal_gyrus | 4.688 | 4.688 | 1 | 28.542 | 5.437 | 0.027 |
| Lingual_gyrus | 0.031 | 0.031 | 1 | 30.351 | 0.135 | 0.715 |
| Cuneus | 0.240 | 0.240 | 1 | 32.469 | 1.806 | 0.188 |
| Medial_orbital_gyrus | 0.626 | 0.626 | 1 | 19.761 | 18.564 | 0.000 |
| Lateral_orbital_gyrus | 0.123 | 0.123 | 1 | 31.346 | 12.030 | 0.002 |

| Regions | Sum Sq | Mean Sq | NumDF | DenDF | F value | Pr(>F) |
|---|---|---|---|---|---|---|
| Posterior_orbital_gyrus | 1.216 | 1.216 | 1 | 30.105 | 30.782 | 0.000 |
| Substantia_nigra | 0.000 | 0.000 | 1 | 35.502 | 0.039 | 0.845 |
| Subgenual_frontal_cortex | 0.046 | 0.046 | 1 | 10.831 | 20.125 | 0.001 |
| Subcallosal_area | 0.001 | 0.001 | 1 | 25.010 | 6.088 | 0.021 |
| Presubgenual_frontal_cortex | 0.037 | 0.037 | 1 | 18.396 | 24.541 | 0.000 |
| Superior_temporal_gyrus_anterior_part | 0.366 | 0.366 | 1 | 26.688 | 28.540 | 0.000 |
| Cerebellum_gm | 72.391 | 72.391 | 1 | 16.329 | 21.391 | 0.000 |
| Cerebellum_wm | 0.672 | 0.672 | 1 | 29.572 | 0.540 | 0.468 |
| Cerebellum_dentate | 0.034 | 0.034 | 1 | 36.300 | 2.708 | 0.108 |

Figure 2

Spaghetti plots for patients across the 43 ROIs showing the estimated annual brain volume loss

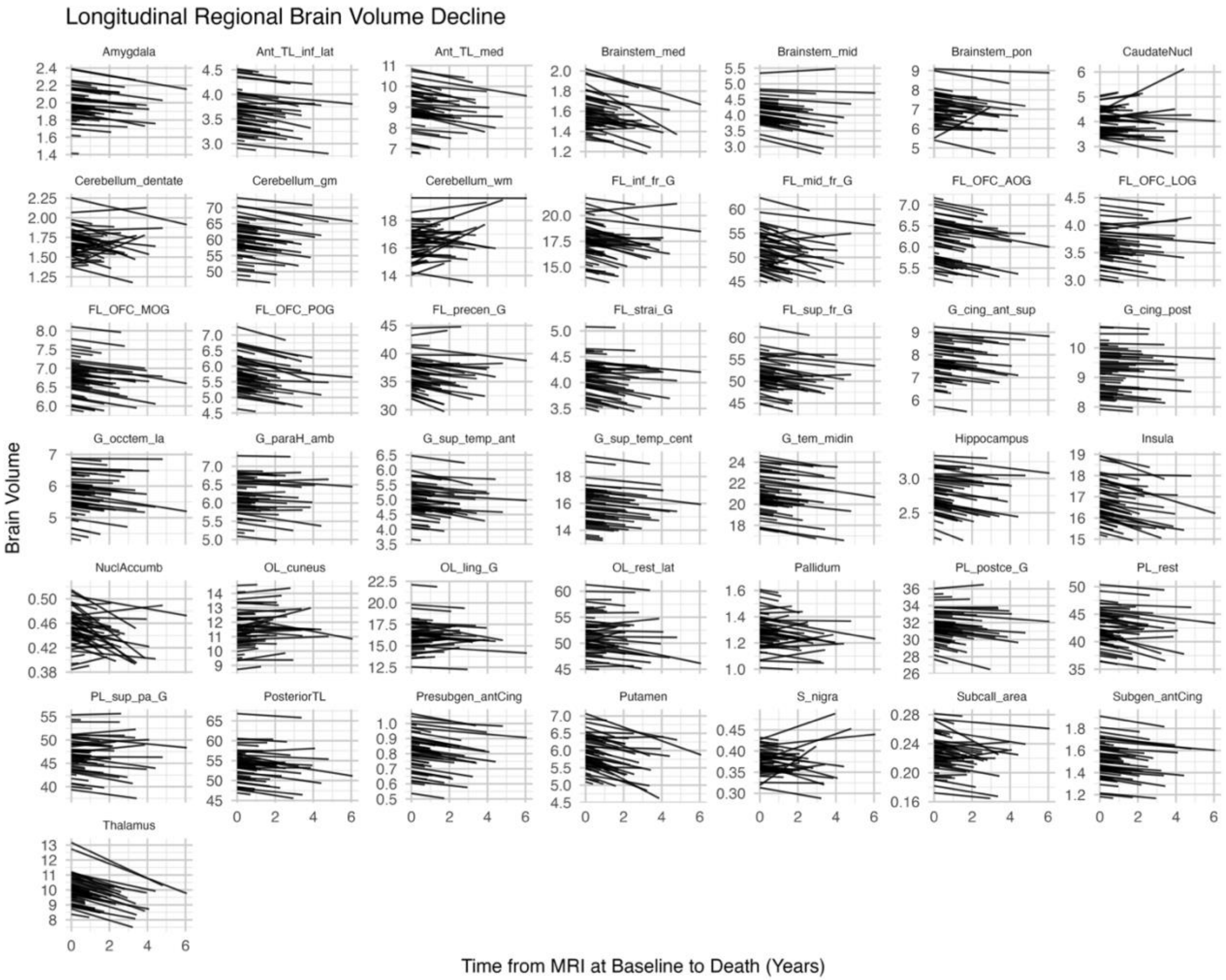

Longitudinal Regional Brain Volume Decline
Amygdala
Ant_TL_inf_lat
Ant_TL_med
Brainstem_med
Brainstem_mid
Brainstem_pon
CaudateNucl
Cerebellum_dentate
Cerebellum_gm
Cerebellum_wm
FL_inf_fr_G
FL_mid_fr_G
FL_OFC_AOG
FL_OFC_LOG
FL_OFC_MOG
FL_OFC_POG
FL_precen_G
FL_strai_G
FL_sup_fr_G
G_cing_ant_sup
G_cing_post
G_occtem_la
G_paraH_amb
G_sup_temp_ant
G_sup_temp_cent
G_tem_midin
Hippocampus
Insula
NuclAccumb
OL_cuneus
OL_ling_G
OL_rest_lat
Pallidum
PL_postce_G
PL_rest
PL_sup_pa_G
PosteriorTL
Presubgen_antCing
Putamen
S_nigra
Subcall_area
Subgen_antCing
Thalamus
Brain Volume
Time from MRI at Baseline to Death (Years)

Table 2

Explorative partial Pearsons correlations (not adjusted for multiple comparisons) using the MRI cohort between MRI at baseline components and survival rate adjusted for disease duration

| Component | r | p-value |
|---|---|---|
| Component 1 | -0.07 | 0.30 |
| Component 3 | -0.24 | 0.03 |
| Component 4 | -0.11 | 0.20 |
| Component 5 | -0.02 | 0.43 |
| Component 6 | -0.20 | 0.05 |
| Component 7 | -0.20 | 0.05 |
| Component 8 | 0.07 | 0.29 |
| Component 9 | 0.23 | 0.03 |
| Component 10 | 0.21 | 0.05 |

Table 3

Explorative partial Pearsons correlations (not adjusted for multiple comparisons) using the MRI cohort between MRI-Slopes components and survival rate adjusted for disease duration

| Component | r | p-value |
|---|---|---|
| Component 1 | -0.25 | 0.02 |
| Component 2 | 0.23 | 0.04 |
| Component 3 | 0.30 | 0.009 |
| Component 4 | 0.16 | 0.11 |
| Component 5 | 0.03 | 0.39 |
| Component 7 | 0.31 | 0.008 |
| Component 8 | -0.13 | 0.15 |
| Component 9 | 0.09 | 0.24 |
| Component 10 | -0.13 | 0.15 |

Table 4

Explorative partial Spearman's correlations (not adjusted for multiple comparisons) using the PET cohort between TSPO PET components and survival rate adjusted for disease duration

| Component | rho | p-value | Bayes Factor |
|---|---|---|---|
| Component 1 | 0.22 | 0.79 | 0.17 |
| Component 3 | 0.04 | 0.56 | 0.22 |
| Component 4 | -0.16 | 0.27 | 0.52 |

Table 5

Explorative partial Spearman's correlations (not adjusted for multiple comparisons) using the PET cohort between MRI-Cross Sectional components and survival rate adjusted for disease duration

| Component | rho | p-value | Bayes Factor |
|---|---|---|---|
| Component 1 | -0.07 | 0.39 | 0.50 |
| Component 3 | -0.38 | 0.06 | 1.46 |
| Component 4 | -0.02 | 0.49 | 0.29 |
| Component 5 | -0.04 | 0.44 | 0.47 |
| Component 6 | 0.07 | 0.61 | 0.03 |

Table 6

Explorative partial Spearman's correlations (not adjusted for multiple comparisons) using the PET cohort between plasma fluid biomarkers and survival adjusted for disease duration

| Biomarker | rho | p-value | Bayes Factor |
|---|---|---|---|
| GFAP | -0.16 | 0.28 | 0.49 |
| AB40 | -0.05 | 0.43 | 0.39 |
| AB42 | -0.22 | 0.22 | 0.56 |
| pTau181 | -0.22 | 0.22 | 0.70 |
| AB42 / AB40 | -0.26 | 0.19 | 0.75 |
| GFAP/NfL | 0.27 | 0.18 | 0.16 |

Table 7
Regional rotated loading of the four $[^{11}C]$-PK11195 PET components derived by principal component analysis on non-displaceable binding potential regional values (PET Cohort).

| Regions | Dim.1 | Dim.2 | Dim.3 | Dim.4 |
|---|---|---|---|---|
| Hippocampus | 0.185 | 0.342 | -0.713 | -0.231 |
| Amygdala | 0.178 | 0.483 | -0.741 | -0.119 |
| Anterior_temporal_lobe_medial_part | 0.313 | 0.083 | -0.911 | 0.016 |
| Anterior_temporal_lobe_lateral_part | 0.621 | 0.151 | -0.704 | -0.133 |
| Parahippocampal_and_ambient_gyri | 0.500 | 0.211 | -0.714 | -0.138 |
| Superior_temporal_gyrus_posterior_part | 0.729 | 0.294 | -0.503 | -0.151 |
| Middle_and_inferior_temporal_gyrus | 0.727 | 0.248 | -0.599 | -0.047 |
| Fusiform_gyrus | 0.575 | 0.241 | -0.513 | 0.010 |
| Insula | 0.540 | 0.400 | -0.672 | -0.159 |
| Lateral_remainder_of_occipital_lobe | 0.674 | 0.261 | -0.596 | -0.216 |
| Cingulate_gyrus_anterior_part | 0.531 | 0.006 | -0.666 | 0.022 |
| Gyrus_cinguli_posterior_part | 0.702 | 0.166 | -0.596 | -0.029 |
| Middle_frontal_gyrus | 0.626 | 0.279 | -0.173 | -0.326 |
| Posterior_temporal_lobe | 0.699 | 0.297 | -0.629 | -0.102 |
| Inferiolateral_remainder_of_parietal_lobe | 0.660 | 0.144 | -0.668 | -0.160 |
| Caudate_nucleus | 0.681 | 0.162 | -0.349 | 0.458 |

| Regions | Dim.1 | Dim.2 | Dim.3 | Dim.4 |
|---|---|---|---|---|
| Nucleus_accumbens | 0.406 | -0.099 | -0.509 | 0.466 |
| Putamen | 0.540 | 0.355 | -0.253 | 0.159 |
| Thalamus | 0.857 | 0.266 | -0.203 | -0.014 |
| Pallidum | 0.012 | 0.732 | -0.149 | 0.118 |
| Precentral_gyrus | 0.393 | 0.443 | -0.372 | -0.592 |
| Straight_gyrus | 0.759 | 0.087 | -0.564 | 0.168 |
| Anterior_orbital_gyrus | 0.844 | 0.267 | -0.328 | 0.058 |
| Inferior_frontal_gyrus | 0.829 | 0.146 | -0.451 | -0.090 |
| Superior_frontal_gyrus | 0.721 | 0.095 | -0.039 | -0.234 |
| Postcentral_gyrus | 0.547 | 0.161 | -0.583 | -0.332 |
| Superior_parietal_gyrus | 0.562 | 0.046 | -0.677 | -0.343 |
| Lingual_gyrus | 0.799 | 0.140 | -0.497 | 0.017 |
| Cuneus | 0.802 | 0.123 | -0.439 | -0.022 |
| Medial_orbital_gyrus | 0.847 | 0.211 | -0.409 | 0.096 |
| Lateral_orbital_gyrus | 0.795 | 0.050 | -0.505 | -0.037 |
| Posterior_orbital_gyrus | 0.644 | 0.271 | -0.667 | -0.054 |
| Substantia_nigra | -0.047 | 0.028 | -0.110 | -0.715 |
| Subgenual_frontal_cortex | 0.691 | 0.349 | -0.456 | 0.215 |

| Regions | Dim.1 | Dim.2 | Dim.3 | Dim.4 |
|---|---|---|---|---|
| Subcallosal_area | 0.348 | 0.096 | -0.732 | 0.505 |
| Presubgenual_frontal_cortex | 0.841 | 0.142 | -0.293 | 0.098 |
| Superior_temporal_gyrus_anterior_part | 0.538 | 0.068 | -0.781 | 0.216 |
| Cerebellum_gm | 0.850 | 0.019 | -0.320 | 0.239 |
| Cerebellum_wm | 0.093 | 0.798 | -0.352 | -0.116 |
| Cerebellum_dentate | 0.221 | 0.813 | 0.044 | -0.058 |
| Brainstem_med | 0.177 | 0.495 | -0.504 | -0.440 |
| Brainstem_mid | 0.645 | 0.526 | -0.175 | -0.335 |
| Brainstem_pon | 0.355 | 0.829 | -0.057 | -0.123 |
| **Cumulative % of var.** | **64** | **72** | **78** | **82** |

Table 8
Regional rotated loading of the 6 components derived by principal component analysis on MRI Cross Sectional (PET Cohort).

| Regions | Dim.1 | Dim.2 | Dim.3 | Dim.4 | Dim.5 | Dim.6 |
|---|---|---|---|---|---|---|
| Brainstem_mid | -0.730 | -0.598 | 0.181 | -0.034 | -0.056 | -0.050 |
| Brainstem_pon | -0.458 | -0.827 | -0.090 | 0.052 | 0.107 | -0.061 |
| Brainstem_med | -0.241 | -0.833 | -0.125 | -0.133 | -0.041 | -0.273 |
| Hippocampus | -0.179 | -0.559 | -0.702 | -0.019 | 0.088 | 0.092 |
| Amygdala | 0.122 | -0.521 | -0.692 | 0.057 | 0.004 | 0.086 |
| Anterior_temporal_lobe_medial_part | 0.521 | -0.250 | -0.718 | 0.086 | -0.135 | -0.029 |
| Anterior_temporal_lobe_lateral_part | 0.445 | -0.533 | -0.375 | -0.148 | 0.019 | -0.264 |
| Parahippocampal_and_ambient_gyri | 0.214 | -0.257 | -0.419 | 0.406 | 0.164 | 0.269 |
| Superior_temporal_gyrus_posterior_part | -0.291 | 0.039 | -0.785 | -0.155 | 0.132 | -0.187 |
| Middle_and_inferior_temporal_gyrus | -0.106 | 0.099 | -0.726 | -0.284 | 0.036 | -0.309 |
| Fusiform_gyrus | 0.274 | 0.417 | -0.688 | -0.214 | 0.248 | -0.149 |
| Insula | -0.723 | 0.231 | -0.352 | -0.158 | -0.021 | 0.072 |
| Lateral_remainder_of_occipital_lobe | -0.673 | 0.532 | 0.061 | -0.060 | -0.184 | -0.181 |
| Cingulate_gyrus_anterior_part | -0.751 | -0.003 | -0.218 | 0.307 | 0.259 | 0.018 |
| Gyrus_cinguli_posterior_part | -0.801 | 0.391 | 0.159 | 0.052 | 0.203 | -0.038 |
| Middle_frontal_gyrus | -0.911 | 0.071 | -0.287 | 0.162 | 0.038 | -0.084 |

| Regions | Dim.1 | Dim.2 | Dim.3 | Dim.4 | Dim.5 | Dim.6 |
|---|---|---|---|---|---|---|
| Posterior_temporal_lobe | -0.498 | 0.380 | -0.471 | -0.293 | 0.252 | -0.240 |
| Inferiolateral_remainder_of_parietal_lobe | -0.687 | 0.332 | -0.082 | 0.025 | 0.407 | -0.148 |
| Caudate_nucleus | -0.784 | 0.050 | -0.001 | 0.040 | -0.435 | -0.051 |
| Nucleus_accumbens | -0.478 | -0.303 | -0.015 | -0.605 | -0.401 | -0.108 |
| Putamen | -0.819 | -0.278 | -0.294 | -0.085 | -0.039 | 0.072 |
| Thalamus | -0.569 | -0.609 | 0.154 | 0.132 | -0.116 | -0.050 |
| Pallidum | -0.760 | -0.238 | 0.134 | -0.264 | -0.434 | -0.160 |
| Precentral_gyrus | -0.692 | 0.104 | -0.280 | 0.496 | 0.255 | -0.092 |
| Straight_gyrus | -0.222 | 0.516 | -0.134 | -0.469 | -0.107 | 0.267 |
| Anterior_orbital_gyrus | -0.576 | 0.098 | -0.208 | 0.352 | -0.371 | 0.291 |
| Inferior_frontal_gyrus | -0.787 | 0.082 | 0.019 | -0.195 | 0.174 | 0.456 |
| Superior_frontal_gyrus | -0.774 | 0.290 | -0.232 | 0.373 | -0.028 | -0.047 |
| Postcentral_gyrus | -0.619 | 0.242 | -0.064 | 0.598 | 0.014 | 0.142 |
| Superior_parietal_gyrus | -0.800 | 0.244 | 0.237 | 0.246 | 0.185 | 0.095 |
| Lingual_gyrus | -0.360 | 0.267 | 0.073 | 0.051 | -0.677 | -0.342 |
| Cuneus | -0.381 | 0.417 | -0.171 | 0.305 | -0.233 | -0.552 |
| Medial_orbital_gyrus | -0.735 | 0.373 | -0.226 | 0.120 | -0.180 | 0.267 |
| Lateral_orbital_gyrus | -0.381 | -0.126 | -0.169 | -0.044 | -0.537 | 0.313 |

| Regions | Dim.1 | Dim.2 | Dim.3 | Dim.4 | Dim.5 | Dim.6 |
|---|---|---|---|---|---|---|
| Posterior_orbital_gyrus | -0.149 | 0.154 | -0.513 | -0.357 | -0.174 | 0.612 |
| Substantia_nigra | -0.759 | -0.371 | 0.232 | 0.114 | -0.004 | -0.118 |
| Subgenual_frontal_cortex | -0.676 | 0.181 | 0.081 | -0.633 | 0.193 | -0.033 |
| Subcallosal_area | -0.384 | -0.127 | 0.234 | -0.563 | -0.019 | 0.087 |
| Presubgenual_frontal_cortex | -0.567 | -0.038 | 0.107 | -0.527 | 0.463 | 0.018 |
| Superior_temporal_gyrus_anterior_part | 0.581 | -0.206 | -0.607 | 0.174 | -0.268 | 0.109 |
| Cerebellum_gm | -0.293 | -0.806 | 0.028 | 0.034 | 0.019 | 0.013 |
| Cerebellum_wm | -0.545 | -0.717 | 0.215 | 0.027 | 0.166 | 0.139 |
| Cerebellum_dentate | -0.608 | -0.623 | 0.285 | 0.128 | 0.230 | -0.011 |
| **Cumulative % of var.** | **33** | **50** | **62** | **70** | **76** | **81** |

Table 9
Regional rotated loading of the 10 components derived by principal component analysis on MRI-Slopes (MRI Cohort).

| Regions | Dim.1 | Dim.2 | Dim.3 | Dim.4 | Dim.5 | Dim.6 | Dim.7 | Dim.8 | Dim.9 | Dim.10 |
|---|---|---|---|---|---|---|---|---|---|---|
| Brainstem_mid | 0.164 | -0.225 | -0.171 | 0.095 | -0.103 | -0.084 | -0.500 | 0.516 | -0.024 | 0.158 |
| Brainstem_pon | 0.068 | 0.153 | -0.111 | 0.114 | 0.086 | 0.840 | -0.194 | 0.092 | -0.141 | 0.077 |
| Brainstem_med | -0.183 | -0.156 | 0.100 | 0.149 | 0.010 | 0.804 | 0.291 | -0.014 | -0.137 | 0.123 |
| Hippocampus | 0.166 | -0.185 | -0.205 | -0.680 | -0.076 | -0.138 | -0.224 | 0.049 | -0.101 | 0.068 |
| Amygdala | 0.171 | 0.177 | -0.007 | -0.049 | -0.262 | 0.529 | 0.253 | 0.178 | 0.373 | 0.235 |
| Anterior_temporal_lobe_medial_part | 0.272 | 0.146 | 0.021 | -0.538 | -0.367 | 0.315 | -0.083 | 0.327 | 0.072 | 0.002 |
| Anterior_temporal_lobe_lateral_part | 0.292 | -0.026 | -0.055 | -0.326 | -0.682 | 0.066 | 0.112 | 0.253 | 0.273 | 0.217 |
| Parahippocampal_and_ambient_gyri | -0.060 | 0.019 | -0.018 | -0.717 | -0.023 | -0.131 | -0.005 | -0.144 | -0.346 | -0.132 |
| Superior_temporal_gyrus_posterior_part | 0.277 | 0.013 | 0.316 | -0.290 | -0.142 | 0.266 | 0.161 | 0.451 | -0.250 | 0.012 |
| Middle_and_inferior_temporal_gyrus | 0.225 | -0.106 | 0.314 | -0.676 | -0.285 | -0.123 | 0.101 | 0.215 | -0.059 | -0.003 |
| Fusiform_gyrus | 0.158 | 0.013 | 0.116 | -0.799 | 0.060 | 0.016 | 0.079 | -0.138 | 0.078 | 0.060 |
| Insula | 0.637 | -0.064 | 0.167 | -0.418 | -0.128 | 0.218 | 0.016 | 0.422 | -0.193 | -0.012 |
| Lateral_remainder_of_occipital_lobe | 0.086 | -0.107 | 0.852 | -0.123 | -0.101 | -0.104 | 0.014 | -0.128 | -0.077 | 0.019 |
| Cingulate_gyrus_anterior_part | 0.755 | -0.420 | -0.079 | -0.076 | -0.216 | 0.068 | -0.146 | 0.009 | 0.081 | 0.088 |
| Gyrus_cinguli_posterior_part | 0.123 | -0.854 | 0.091 | 0.027 | -0.103 | 0.101 | -0.060 | -0.078 | 0.071 | 0.107 |
| Middle_frontal_gyrus | 0.869 | -0.191 | 0.208 | 0.128 | -0.172 | -0.026 | 0.062 | 0.054 | -0.004 | 0.008 |

| Regions | Dim.1 | Dim.2 | Dim.3 | Dim.4 | Dim.5 | Dim.6 | Dim.7 | Dim.8 | Dim.9 | Dim.10 |
|---|---|---|---|---|---|---|---|---|---|---|
| Posterior_temporal_lobe | 0.116 | -0.294 | 0.571 | -0.509 | -0.290 | -0.100 | 0.092 | 0.012 | -0.167 | 0.011 |
| Inferiolateral_remainder_of_parietal_lobe | 0.402 | -0.603 | 0.506 | -0.143 | -0.088 | 0.001 | 0.020 | -0.007 | -0.211 | 0.038 |
| Caudate_nucleus | 0.170 | 0.024 | -0.089 | 0.223 | -0.115 | -0.012 | 0.033 | 0.773 | -0.020 | -0.026 |
| Nucleus_accumbens | 0.181 | 0.214 | 0.239 | -0.045 | -0.195 | 0.116 | -0.266 | 0.096 | 0.398 | 0.586 |
| Putamen | 0.280 | -0.055 | -0.096 | -0.305 | -0.101 | 0.283 | -0.059 | 0.679 | 0.181 | 0.199 |
| Thalamus | -0.095 | 0.121 | 0.184 | 0.085 | -0.315 | 0.116 | 0.778 | 0.085 | 0.043 | 0.180 |
| Pallidum | -0.010 | 0.268 | 0.162 | 0.111 | 0.106 | 0.372 | -0.011 | 0.123 | -0.681 | -0.326 |
| Precentral_gyrus | 0.450 | -0.659 | 0.025 | -0.033 | 0.102 | -0.018 | -0.150 | 0.143 | 0.143 | -0.265 |
| Straight_gyrus | 0.479 | -0.282 | -0.146 | -0.297 | 0.214 | -0.085 | 0.470 | 0.038 | 0.033 | -0.056 |
| Anterior_orbital_gyrus | 0.857 | -0.160 | 0.008 | -0.293 | -0.076 | 0.069 | -0.088 | 0.006 | -0.039 | -0.047 |
| Inferior_frontal_gyrus | 0.837 | -0.211 | -0.100 | -0.064 | -0.193 | 0.068 | 0.057 | 0.191 | 0.200 | -0.017 |
| Superior_frontal_gyrus | 0.868 | -0.293 | 0.107 | -0.022 | -0.136 | 0.022 | -0.009 | 0.066 | 0.127 | 0.017 |
| Postcentral_gyrus | 0.157 | -0.677 | -0.059 | -0.186 | 0.102 | -0.171 | 0.029 | 0.171 | 0.102 | -0.127 |
| Superior_parietal_gyrus | 0.311 | -0.776 | 0.367 | -0.035 | 0.006 | -0.135 | 0.024 | -0.205 | -0.013 | -0.103 |
| Lingual_gyrus | -0.258 | -0.041 | 0.695 | 0.185 | -0.114 | 0.195 | 0.102 | 0.012 | -0.255 | 0.230 |
| Cuneus | 0.047 | -0.059 | 0.922 | -0.008 | 0.233 | -0.002 | 0.078 | -0.015 | -0.021 | -0.010 |
| Medial_orbital_gyrus | 0.662 | -0.247 | -0.044 | -0.421 | 0.325 | -0.113 | 0.276 | 0.018 | 0.071 | -0.081 |
| Lateral_orbital_gyrus | 0.776 | -0.296 | 0.016 | -0.327 | 0.019 | -0.181 | -0.071 | 0.029 | 0.036 | 0.077 |

| Regions | Dim.1 | Dim.2 | Dim.3 | Dim.4 | Dim.5 | Dim.6 | Dim.7 | Dim.8 | Dim.9 | Dim.10 |
|---|---|---|---|---|---|---|---|---|---|---|
| Posterior_orbital_gyrus | 0.701 | 0.320 | -0.104 | -0.114 | -0.223 | 0.014 | -0.180 | 0.237 | 0.118 | -0.168 |
| Substantia_nigra | 0.216 | 0.398 | -0.221 | -0.145 | 0.081 | 0.447 | -0.442 | 0.059 | -0.365 | 0.140 |
| Subgenual_frontal_cortex | 0.621 | 0.243 | 0.093 | 0.089 | 0.075 | 0.048 | 0.342 | 0.465 | 0.095 | 0.142 |
| Subcallosal_area | 0.260 | 0.255 | -0.083 | -0.145 | 0.073 | -0.001 | 0.266 | 0.389 | -0.019 | 0.577 |
| Presubgenual_frontal_cortex | 0.828 | 0.134 | 0.000 | -0.053 | 0.019 | -0.027 | -0.124 | 0.335 | 0.135 | 0.148 |
| Superior_temporal_gyrus_anterior_part | 0.454 | -0.038 | 0.061 | -0.122 | -0.752 | -0.126 | 0.142 | 0.121 | 0.107 | 0.163 |
| Cerebellum_gm | -0.184 | -0.045 | 0.137 | 0.106 | -0.253 | 0.345 | 0.076 | 0.004 | -0.095 | 0.725 |
| Cerebellum_wm | -0.147 | 0.096 | 0.160 | -0.148 | 0.002 | 0.054 | -0.100 | -0.179 | -0.842 | 0.203 |
| Cerebellum_dentate | -0.178 | 0.001 | 0.144 | -0.182 | 0.111 | 0.044 | 0.009 | 0.100 | -0.833 | -0.046 |
| **Cumulative % of var.** | **26** | **38** | **48** | **56** | **63** | **68** | **72** | **75** | **78** | **80** |

Table 10
Regional rotated loading of the 10 components derived by principal component analysis on MRI at Baseline (MRI Cohort).

| Regions | Dim.1 | Dim.2 | Dim.3 | Dim.4 | Dim.5 | Dim.6 | Dim.7 | Dim.8 | Dim.9 | Dim.10 |
|---|---|---|---|---|---|---|---|---|---|---|
| Brainstem_mid | -0.064 | -0.804 | 0.176 | 0.063 | 0.096 | 0.085 | -0.170 | -0.176 | -0.099 | 0.152 |
| Brainstem_pon | -0.208 | -0.826 | -0.161 | -0.022 | 0.152 | -0.105 | -0.012 | -0.177 | -0.021 | -0.070 |
| Brainstem_med | -0.040 | -0.740 | -0.216 | -0.077 | 0.202 | -0.082 | -0.079 | -0.290 | -0.003 | 0.231 |
| Hippocampus | -0.175 | -0.247 | -0.670 | -0.178 | 0.339 | -0.088 | -0.182 | 0.073 | -0.135 | 0.316 |
| Amygdala | -0.012 | -0.222 | -0.682 | -0.151 | 0.451 | -0.225 | -0.091 | 0.059 | -0.007 | 0.041 |
| Anterior_temporal_lobe_medial_part | -0.022 | -0.017 | -0.931 | -0.052 | 0.008 | -0.028 | -0.131 | -0.061 | 0.039 | -0.009 |
| Anterior_temporal_lobe_lateral_part | 0.069 | -0.103 | -0.534 | -0.151 | 0.101 | -0.029 | -0.231 | -0.584 | 0.150 | 0.025 |
| Parahippocampal_and_ambient_gyri | -0.108 | -0.223 | -0.634 | -0.284 | 0.081 | 0.179 | -0.139 | 0.138 | -0.242 | 0.129 |
| Superior_temporal_gyrus_posterior_part | -0.450 | -0.060 | -0.505 | 0.017 | -0.180 | -0.437 | 0.009 | -0.224 | -0.121 | -0.008 |
| Middle_and_inferior_temporal_gyrus | -0.338 | 0.005 | -0.600 | -0.165 | 0.086 | -0.283 | -0.159 | -0.349 | -0.232 | -0.127 |
| Fusiform_gyrus | -0.373 | 0.152 | -0.695 | 0.042 | -0.237 | -0.064 | 0.174 | 0.022 | -0.210 | -0.013 |
| Insula | -0.503 | -0.163 | -0.191 | -0.156 | 0.377 | -0.221 | -0.062 | -0.005 | -0.435 | 0.235 |
| Lateral_remainder_of_occipital_lobe | -0.351 | -0.042 | -0.129 | -0.714 | 0.179 | -0.231 | -0.247 | 0.012 | -0.263 | 0.019 |
| Cingulate_gyrus_anterior_part | -0.595 | -0.131 | -0.208 | 0.028 | 0.162 | -0.351 | -0.322 | 0.361 | -0.032 | 0.009 |
| Gyrus_cinguli_posterior_part | -0.747 | -0.077 | 0.090 | -0.160 | 0.061 | -0.207 | -0.276 | 0.176 | -0.124 | 0.058 |
| Middle_frontal_gyrus | -0.764 | -0.098 | -0.097 | -0.104 | 0.264 | -0.195 | -0.343 | -0.055 | 0.001 | 0.148 |

| Regions | Dim.1 | Dim.2 | Dim.3 | Dim.4 | Dim.5 | Dim.6 | Dim.7 | Dim.8 | Dim.9 | Dim.10 |
|---|---|---|---|---|---|---|---|---|---|---|
| Posterior_temporal_lobe | -0.610 | -0.068 | -0.393 | -0.164 | 0.016 | -0.396 | 0.036 | -0.062 | -0.371 | 0.015 |
| Inferiolateral_remainder_of_parietal_lobe | -0.793 | -0.093 | -0.194 | -0.027 | 0.012 | -0.312 | 0.013 | -0.182 | -0.174 | 0.186 |
| Caudate_nucleus | -0.108 | -0.182 | 0.080 | -0.155 | 0.775 | 0.196 | 0.047 | -0.002 | -0.175 | 0.054 |
| Nucleus_accumbens | -0.038 | 0.035 | -0.091 | -0.264 | 0.681 | -0.369 | -0.141 | -0.142 | 0.091 | 0.088 |
| Putamen | -0.200 | -0.290 | -0.036 | -0.036 | 0.813 | -0.270 | -0.031 | 0.048 | -0.066 | 0.049 |
| Thalamus | -0.258 | -0.335 | -0.129 | 0.022 | 0.180 | -0.114 | -0.072 | 0.006 | -0.022 | 0.797 |
| Pallidum | -0.139 | -0.494 | 0.060 | -0.249 | 0.248 | 0.060 | -0.011 | -0.058 | -0.614 | 0.152 |
| Precentral_gyrus | -0.776 | -0.415 | -0.052 | -0.081 | 0.161 | 0.090 | -0.062 | 0.031 | -0.012 | 0.074 |
| Straight_gyrus | -0.206 | 0.107 | -0.141 | -0.052 | 0.011 | -0.272 | -0.407 | 0.111 | -0.677 | -0.133 |
| Anterior_orbital_gyrus | -0.396 | 0.033 | -0.152 | -0.321 | 0.128 | 0.020 | -0.649 | -0.068 | -0.042 | 0.097 |
| Inferior_frontal_gyrus | -0.480 | -0.121 | -0.057 | -0.087 | 0.122 | -0.212 | -0.413 | -0.052 | -0.254 | 0.357 |
| Superior_frontal_gyrus | -0.777 | -0.063 | -0.059 | -0.047 | -0.036 | -0.146 | -0.274 | 0.065 | -0.053 | -0.025 |
| Postcentral_gyrus | -0.779 | -0.260 | -0.093 | -0.022 | 0.167 | 0.167 | -0.099 | -0.033 | -0.026 | -0.156 |
| Superior_parietal_gyrus | -0.679 | -0.113 | 0.120 | -0.384 | -0.105 | -0.100 | -0.204 | -0.049 | -0.031 | 0.197 |
| Lingual_gyrus | -0.054 | -0.202 | -0.156 | -0.821 | 0.105 | -0.150 | -0.061 | 0.025 | -0.141 | -0.045 |
| Cuneus | -0.139 | -0.019 | -0.183 | -0.841 | 0.165 | 0.018 | -0.124 | -0.094 | 0.060 | 0.012 |
| Medial_orbital_gyrus | -0.403 | 0.071 | -0.118 | -0.223 | 0.068 | -0.080 | -0.575 | 0.076 | -0.433 | 0.127 |
| Lateral_orbital_gyrus | -0.371 | 0.005 | -0.300 | -0.077 | 0.172 | -0.041 | -0.683 | -0.062 | -0.066 | 0.172 |

| Regions | Dim.1 | Dim.2 | Dim.3 | Dim.4 | Dim.5 | Dim.6 | Dim.7 | Dim.8 | Dim.9 | Dim.10 |
|---|---|---|---|---|---|---|---|---|---|---|
| Posterior_orbital_gyrus | -0.214 | -0.029 | -0.168 | -0.074 | -0.149 | -0.203 | -0.765 | -0.081 | -0.045 | -0.127 |
| Substantia_nigra | -0.247 | -0.767 | 0.143 | -0.196 | 0.117 | -0.041 | -0.100 | -0.139 | -0.146 | -0.375 |
| Subgenual_frontal_cortex | -0.439 | -0.130 | -0.004 | -0.178 | 0.162 | -0.644 | -0.373 | 0.010 | -0.226 | -0.036 |
| Subcallosal_area | -0.138 | -0.189 | -0.094 | -0.234 | 0.191 | -0.668 | -0.003 | -0.073 | -0.023 | 0.249 |
| Presubgenual_frontal_cortex | -0.374 | -0.198 | -0.065 | 0.027 | 0.108 | -0.663 | -0.328 | 0.176 | -0.106 | -0.054 |
| Superior_temporal_gyrus_anterior_part | 0.087 | -0.084 | -0.782 | -0.059 | -0.104 | -0.009 | -0.172 | -0.032 | 0.121 | 0.038 |
| Cerebellum_gm | 0.016 | -0.723 | -0.253 | -0.123 | -0.001 | -0.280 | 0.003 | 0.333 | 0.067 | 0.172 |
| Cerebellum_wm | -0.126 | -0.851 | -0.115 | -0.132 | 0.025 | -0.131 | 0.138 | 0.287 | 0.038 | 0.057 |
| Cerebellum_dentate | -0.246 | -0.806 | -0.141 | 0.022 | 0.026 | -0.069 | 0.127 | 0.199 | 0.001 | 0.150 |
| **Cumulative % of var.** | **34** | **45** | **54** | **60** | **64** | **68** | **72** | **76** | **77** | **80** |

Table 11: MRI manufacturers by models and scanning parameters.

| Manufacturer | Siemens | General Electric |
|---|---|---|
| Model | TrioTim<br>Verio<br>Skyra<br>Prisma | Signa* |
| Slice and thickness | 256 of 1.00 mm thickness<br>256 of 1.10 mm thickness<br>256 of 1.20 mm thickness<br>192 of 1.25 mm thickness | 512 of 1.00 mm thickness |
| Echo time (s) | 0.00286<br>0.00298<br>0.00293<br>0.00288<br>0.00287<br>0.00285<br>0.00292 | 0.003432<br>0.003604 |
| Repetition Time (s) | 2.3<br>2.0 | 0.008876<br>0.0092 |
| Acquisition matrix | 144 x 192<br>176 x 240<br>208 x 256 | 192x512 |
| Voxel size (mm3) | 1.25 x 1.25 x 1.25<br>1 x 1 x 1<br>1.1 x 1.1 x 1.1<br>1.2 x 1 x 1 | 1 x 0.5469 x 0.5469 |
